# Phase I Clinical Trial of Allogeneic Cytomegalovirus-specific T Cells in Combination with Pembrolizumab for Recurrent Glioblastoma

**DOI:** 10.64898/2026.11.17.26367100

**Authors:** David Walker, Paulo Martins, Panteha Khaledi, Hamish Alexander, Adam Stirling, Michelle A. Neller, Jessica Kelly, Vivien Biggs, Hilary Reddiex, Archana Panikkar, Leone Beagley, Shannon Flynn, Jyothy Raju, Meggie Voogt, Matthew Solomon, Corey Smith, Rajiv Khanna

## Abstract

**Purpose:** Glioblastoma (GBM) remains refractory to immunotherapy due to its immunosuppressive microenvironment. Tumour-infiltrating cytomegalovirus (CMV)-specific T cells express elevated programmed cell death protein-1 (PD-1), suggesting checkpoint-mediated dysfunction may limit T-cell therapy. We evaluated the safety and immunological activity of allogeneic CMV-specific T-cell therapy (CYT-101) combined with pembrolizumab.

**Patients and Methods:** In this phase I 3+3 dose-escalation study, recurrent GBM patients received four weekly infusions of CYT-101 followed by pembrolizumab. The primary endpoint was safety. Secondary endpoints included progression-free survival at 6 months (PFS-6), overall survival at 6 months (OS-6), and immune correlates of response.

**Results:** CYT-101 T-cell therapy demonstrated polyfunctional activity, broad HLA restriction, potent cytolytic function, and diverse T-cell receptor repertoires. Nine patients received study treatment. No dose-limiting toxicities were observed, and the maximum tolerated dose was not reached. Only six adverse events were considered treatment-related, and no treatment-related serious adverse events, graft-versus-host disease, cytokine release syndrome, or neurotoxicity occurred. OS-6 and PFS-6 were 55% and 11%, respectively, with a median overall survival of 9.1 months. No objective clinical responses were seen. Of the nine treated participants, one had stable disease, five had progressive disease, and three were unevaluable by modified RANO criteria. Immune profiling revealed enhanced global CD4^+^ and CD8^+^ T-cell function following CYT-101 therapy, while donor-derived CYT-101 clonotypes were detected in six of nine participants, with significant T-cell repertoire remodelling.

**Conclusions:** Allogeneic CMV-specific T-cell therapy combined with pembrolizumab was safe in recurrent GBM, supporting further evaluation of this strategy to overcome immune dysfunction in GBM.

**Statement of Translational Relevance:** Glioblastoma (GBM) remains a major therapeutic challenge, with limited benefit from immune checkpoint blockade or adoptive cellular therapies when used independently. This phase I study provides clinical evidence that combining an off-the-shelf, allogeneic cytomegalovirus (CMV)-specific T-cell therapy (CYT-101) with PD-1 blockade is safe, feasible, and immunologically active in recurrent GBM. CYT-101 demonstrated a favourable safety profile without graft-versus-host disease, cytokine release syndrome, neurotoxicity, or treatment-related serious adverse events. Importantly, donor-derived CYT-101 clonotypes were detected following infusion, accompanied by enhanced systemic CD4^+^ and CD8^+^ T-cell function and substantial T-cell repertoire remodelling following CYT-101 and pembrolizumab. These findings provide proof-of-concept that allogeneic virus-specific T cells can be safely combined with checkpoint inhibition in GBM, supporting further clinical development and evaluation in larger studies.

## Introduction

Glioblastoma (GBM) remains one of the most aggressive and lethal human malignancies (1). Despite advances in neurosurgical techniques, radiotherapy, and chemotherapy, outcomes have improved only marginally over the past three decades. Brain cancer remains the leading cause of cancer-related death in Australians under 40 years of age. For patients with recurrent GBM, the current standard of care provides a median overall survival (OS) of 4.1 months, and median progression-free survival (PFS) of 2.1 months (2). These poor outcomes highlight the urgent need for novel therapeutic approaches.

The limited efficacy of immunotherapy in GBM is largely attributed to its profoundly immunosuppressive tumour microenvironment (3). GBM is generally considered an immunologically “cold” tumour, characterised by low T-cell infiltration, extensive myeloid-mediated immune suppression, T-cell dysfunction, and multiple mechanisms of immune evasion (1,4). Consequently, strategies that simultaneously increase tumour-reactive T-cell numbers and overcome inhibitory immune pathways may be required to achieve meaningful clinical benefit (5,6). Programmed cell death protein 1 (PD-1) is an inhibitory immune checkpoint receptor expressed on activated T cells. Engagement of PD-1 by its ligands, programmed death-ligand 1 (PD-L1) and programmed death-ligand 2 (PD-L2), suppresses T-cell activation and effector function, thereby maintaining peripheral immune tolerance (7,8). Many cancers exploit this pathway to evade immune surveillance, making PD-1 blockade an attractive therapeutic strategy. Pembrolizumab is a humanised immunoglobulin G4 monoclonal antibody that binds PD-1 and prevents its interaction with PD-L1 and PD-L2. Although PD-1 blockade has demonstrated substantial clinical benefit across multiple malignancies, its efficacy in GBM has been limited. Clinical studies of pembrolizumab in recurrent GBM following standard chemoradiotherapy have failed to demonstrate a significant survival benefit (9).

Adoptive cellular immunotherapy using cytomegalovirus (CMV)-specific T cells has emerged as a promising therapeutic approach for GBM (10,11). Multiple studies have reported the expression of CMV antigens within malignant glioma cells, providing a rationale for targeting CMV-associated epitopes in GBM (12–14). Several approaches have been assessed targeting CMV in GBM. These include pp65 mRNA-loaded dendritic cell vaccines and CMV peptide vaccines, and antiviral therapy with valganciclovir (13,15–21). In our previous clinical studies, adoptively transferred CMV-specific T cells trafficked to tumour sites and mediated anti-tumour immune responses (22,23). However, detailed immune profiling revealed that tumour-infiltrating CMV-specific T cells expressed elevated levels of inhibitory checkpoint molecules, including PD-1, compared with their circulating counterparts (23). These findings suggested that transferred CMV-specific T cells may become functionally restrained within the tumour microenvironment through PD-1-mediated immune suppression.

Preclinical and clinical studies across multiple tumour types have demonstrated that immune checkpoint blockade can enhance the expansion, persistence, and effector function of adoptively transferred T cells, supporting a synergistic interaction between adoptive cellular therapy and PD-1 inhibition (24,25). We therefore hypothesised that pembrolizumab could augment the anti-tumour activity of CMV-specific T cells by preventing PD-1-mediated dysfunction within the GBM microenvironment and sustaining T-cell activity at tumour sites. To enable rapid treatment delivery, we developed CYT-101, an allogeneic CMV-specific T-cell therapy generated from healthy CMV-seropositive donors. Compared with autologous manufacturing, this off-the-shelf approach enables immediate treatment availability, reduces the risk of manufacturing failure, minimises product variability, and lowers manufacturing costs. Importantly, virus-specific T cells have demonstrated a favourable safety profile in numerous clinical studies, with minimal risk of graft-versus-host disease owing to their preferential recognition of viral antigens rather than healthy allogeneic tissues.

Here, we report the results of the dose escalation phase of a phase I/II clinical trial, which evaluated the safety of CYT-101 administered in combination with pembrolizumab in patients with recurrent GBM. The primary objective was to determine the maximum tolerated dose and/or recommended dose(s) for future exploration of CYT-101 as a monotherapy or in combination with pembrolizumab. Secondary and exploratory objectives included characterisation of immune responses and evaluation of preliminary signals of clinical activity.

## Patients and Methods

### Regulatory and ethical approvals

This clinical trial (ACTRN12623001126606) was conducted in accordance with the World Medical Association Declaration of Helsinki – Ethical Principles for Medical Research Involving Human Participants, the National Health and Medical Research Council’s National Statement on Ethical Conduct in Human Research (2023), and the Integrated Addendum to ICH E6 (R1): Guideline for Good Clinical Practice E6 (R2) (November 2016) – with introductory comments of the Australian Therapeutic Goods Administration. To generate allogeneic CMV-specific T cell therapies (CYT-101), healthy blood donors were recruited via Stem Cell Donors Australia (formerly the Australian Bone Marrow Donor Registry). Ethical approval for the cell therapy manufacturing project was obtained from the Lifeblood Human Research Ethics Committee (HREC) and the QIMR Berghofer Medical Research Institute HREC. The clinical trial was reviewed and approved by the Metro North Health HREC and the UnitingCare Health HREC. All participants in the manufacturing project and clinical trial provided written informed consent prior to undertaking any study-specific activities.

### Study design

This was phase I of an investigator-initiated, open-label, phase I/II clinical trial for patients with glioblastoma multiforme/astrocytoma grade 4. Phase II of the clinical trial (dose expansion) will be conducted subject to funding. A 3 + 3 design was used for the dose-escalation phase of the study, and patients with recurrent disease were recruited. Following screening, participants received infusions of CYT-101 on Days 0, 7, 14 and 21, with a safety visit on Day 3. The first dose cohort received 2 × 10^7^ cells/infusion, the second received 4 × 10^7^ cells/infusion, and the final dose cohort received 8 × 10^7^ cells/infusion. Subsequently, all participants received pembrolizumab (KEYTRUDA®, Merck Sharp & Dohme [Australia] Pty Limited) on Day 28, followed by safety visits on Days 31, 35 and 49. Participants were monitored for dose-limiting toxicities until Day 49, with data from each dose cohort reviewed by the Data and Safety Monitoring Board once all three participants had reached Day 49. Dose escalation was approved based on accumulating safety data. Participants continued to receive pembrolizumab every 42 days until discontinuation or withdrawal, up to a maximum of 18 infusions. Tumour progression was monitored by magnetic resonance imaging. Scans were evaluated by a radiologist in line with the modified Response Assessment in Neuro-Oncology (mRANO) and immunotherapy RANO criteria. The primary objective of this phase of the study was to determine the maximum-tolerated dose and/or recommended dose(s) for future exploration of allogeneic CMV-specific T cells as monotherapy or in combination with pembrolizumab in this cohort. The secondary objective was to investigate the anti-tumour activity of allogeneic CMV-specific T cells in combination with pembrolizumab.

### Manufacture of CYT-101

Peripheral blood mononuclear cells (PBMC) were isolated from the blood of CMV-seropositive donors selected from the Stem Cell Donors Australia registry. To generate CYT-101, PBMC were mixed at a 7:3 ratio with autologous PBMC pulsed with a pool of peptides from CMV-encoding HLA-restricted T cell epitopes (Supplementary Table 1). PBMC were cultured at 37°C/5% CO_2_/95% RH in G-Rex flasks in TheraPEAK® T-VIVO® Cell Culture Medium (Lonza cat# BP12-970Q) supplemented with 5% PLTGold-GI (Mill Creek Life Sciences cat# PLTGold100GMP) at a cell density of 2 × 10^6^ cells/cm^2^. Cultures were supplemented with 120 IU/mL of recombinant interleukin (IL)-2 (Miltenyi Biotec cat# 170-076-146) on day 3. Cells were reseeded at a density of 1 × 10^6^ cells/cm^2^ every 3 days until harvest. At harvest, cells were washed, counted and cryopreserved at 4 × 10^7^ cells/vial in cryopreservation medium containing CryoStor CS10 (STEMCELL Technologies cat# 07930) and human serum albumin (Octapharma cat# 5400963).

### Assessment of CYT-101 specificity using intracellular cytokine analysis

CYT-101 therapy batches were thawed in Roswell Park Memorial Institute (RPMI) 1640 Medium (Gibco cat# 61870036) supplemented with 10% foetal bovine serum (Hyclone cat# SH30084.03) and 120 IU/mL of IL-2 (Tecin cat# RO 23-6019) and allowed to recover overnight at 37°C/6.5% CO_2_/95% RH. T cells were stimulated with individual HLA-matched peptide epitopes (reconstituted in-house from synthetic CMV peptides purchased from JPT Peptide Technologies GmbH, Berlin, Germany) (Supplementary Table 1) and cultured for 4 hours in the presence of GolgiPlug (BD Biosciences cat# 555028). Cells were then washed and incubated with anti-CD3– APC, anti-CD4–FITC, anti-CD8–PerCP-Cy5.5, and LIVE/DEAD Fixable Near-IR Dead Cell Stain (Thermo Fisher/Invitrogen cat# L34976). Cells were fixed and permeabilised with Cytofix/Cytoperm buffer (BD Biosciences), washed with Perm/Wash buffer (BD Biosciences RRID:AB_2869011), and incubated with anti-IFN-γ–PE. Cells were then acquired on a BD LSRFortessa (BD Biosciences RRID:SCR_018655) with FACSDiva software (BD Biosciences RRID:SCR_001456). T cells were considered specific for an epitope if the response was greater than the mean plus three standard deviations of the response of T cells cultured in the absence of stimulus. A list of antibodies containing RRID numbers can be found in Supplementary Table 2.

### Assessment of T-cell alloreactivity

A panel of K562 (RRID:CVCL_0004) cell lines transfected with a single HLA allele was used to assess potential alloreactivity of CYT-101 batches as previously described (26). A batch was considered alloreactive against an HLA class I allele if >1% of lymphocytes produced IFN-γ following subtraction of the background response generated against untransfected K562 cells.

### T-cell cytotoxicity assay

T-cell cytotoxicity was determined using a flow cytometry-based killing assay. CYT-101 batches and PHA-stimulated T cell (PHA blasts) targets were thawed in RPMI 1640 supplemented with 10% foetal bovine serum and 120 IU/mL of IL-2 and allowed to recover overnight at 37°C/6.5% CO_2_/95% RH. To prepare target cells for the killing assay, PHA blasts were labelled with CellTrace Violet (Thermo Fisher Scientific cat# C34557), then incubated with HLA-matched CMV-encoded peptide epitopes. Epitopes with the same HLA restriction were incubated together. Control PHA blasts were incubated without peptide. PHA-blast target cells were then washed and incubated overnight at an effector-to-target ratio of 10:1 with CYT-101. The following day, cells were washed then incubated with 7-Aminoactinomycin (7-AAD) (BD Biosciences cat# 51-68981A). Cells were then acquired on a BD LSRFortessa with FACSDiva software.

### Single-cell RNA-seq, feature barcode and TCR library preparation

Cryopreserved CYT-101 batches were thawed and treated with 50 nM dasatinib (Sigma-Aldrich / Merck cat# CDS023389) for 30 minutes at 37°C. Cells were counted and viability was assessed using trypan blue exclusion (Sigma-Aldrich / Merck, cat# T8154). Cells were stained with DNA-barcoded dCODE-dextamers (Immundex RRID:SCR_025814) for CMV-reactive TCRs, together with TotalSeq^TM^-C anti-human Hashtag antibodies (BioLegend cat# C0251-4) compatible with the 10x Genomics 5′ Feature Barcode workflow. Following staining, samples were sorted by flow cytometry on FACSAria III Cell Sorter (BD Biosciences RRID:SCR_016695) to enrich for live cells. Up to 50,000 cells were loaded per channel onto a Chromium Controller (10x Genomics, RRID:SCR_019326) using the Chromium Next GEM Single Cell 5′ Gene Expression and V(D)J workflow with Feature Barcode technology (10x Genomics / Millennium Science cat# PN-1000263, PN-1000286, PN-1000190, PN-1000252, PN-1000215, PN-1000250), according to the manufacturer’s instructions. Gel bead-in-emulsions were generated, and reverse transcription was performed to barcode cellular transcripts and antibody/dextramer-derived feature barcode tags. Amplified cDNA was used to generate separate 5′ gene expression, feature barcode and TCR V(D)J libraries. Indexed libraries were quantified, pooled and sequenced on an Illumina Nextseq 2000 sequencing platform (Illumina RRID:SCR_023614).

### Single-cell sequencing data processing and analysis

Raw sequencing data for pre- and post-CYT-101, including gene expression (RNA), V(D)J, and cell surface protein libraries, were processed using the Cell Ranger toolkit v9.0.0 (10x Genomics, RRID:SCR_017344). Cellranger count and cellranger vdj pipelines were run against the GRCh38 reference genome with default settings to generate gene expression matrices, antibody-derived tag (ADT) counts, and filtered V(D)J contig annotations. Processed outputs were imported into R v4.5.0 (R Project for Statistical Computing RRID:SCR_001905) for downstream analysis using Seurat v5.2.0 (Satija Lab / Serrat RRID:SCR_016341).

### Quality control for single-cell data

Gene expression matrices were loaded into Seurat v5.2.0 to generate individual sample objects, with ADT data for barcoded multimers incorporated as a separate assay where applicable. Standard per-cell quality metrics were calculated, including total UMI counts, number of detected genes, and the proportion of mitochondrial, ribosomal, and haemoglobin transcripts. Cells were excluded if they exceeded the following thresholds: mitochondrial transcripts ≥5%, ribosomal transcripts ≥40%, haemoglobin transcripts ≥0.1%, or if they contained fewer than 200 detected genes or 200 UMIs. To reduce bias from highly abundant antigen receptor transcripts, T-cell receptor (TRα/β) and immunoglobulin variable genes were removed from the RNA assay prior to normalisation. Putative doublets were identified using DoubletFinder v2 (DoubletFinder, RRID:SCR:018771), and cell-cycle phase scores (S and G2/M) were calculated for each cell. Data were normalised using SCTransform (sctransform RRID:SCR_022146), with mitochondrial transcript proportion regressed out and 2,000 variable features selected for downstream analyses. Multimer specificity of cells was determined using a CLR-normalised cut-off of 1.0.

### Single-cell TCR sequencing data processing

Filtered V(D)J contig annotations generated by Cell Ranger toolkit v9.0.0 were processed to retain productive TRα and TRβ sequences and assign paired T-cell receptors to individual cells. In cases where multiple TRα or TRβ chains were detected, the contig with the highest UMI count was selected, while ties or entries lacking UMI information were excluded. Cells without a detectable TRβ chain were removed, as clonotypes were defined based on TRβ V gene usage, J gene usage, and CDR3 amino acid sequence. The curated TCR dataset was then integrated with gene expression metadata by matching cell barcodes, ensuring that only cells retained after RNA and ADT quality control were included in downstream analyses.

### Single-cell batch integration and clustering

Processed data were merged and used for dimensionality reduction, clustering and UMAP visualisation. Clusters were annotated using cluster-specific differentially expressed genes.

### TCR beta variable (TRBV) analysis

PBMC were isolated from peripheral blood throughout the course of the study and cryopreserved prior to use. Three time points were selected for TRBV deep sequencing analysis: directly prior to the first CYT-101 infusion, prior to the final CYT-101 infusion, and throughout the course of pembrolizumab infusions. DNA was extracted at each time point from between 9 × 10^5^ and 5 × 10^6^ cells. TRBV deep sequencing analysis performed by ImmunoSEQ® (Adaptive Biotechnologies, RRID:SCR_014709). The TRBV CDR3 dataset was then extracted from the Adaptive Biotechnologies Connector portal.

### Deep-sequencing TCR diversity analysis

TCR repertoire diversity was assessed from deep-sequencing data using Shannon entropy, richness, and clonality. To account for differences in sequencing depth between samples, repertoire metrics were estimated using bootstrap resampling. For each sample, 50,000 productive TCR templates were randomly resampled for 10,000 iterations. Within each bootstrap iteration, clonotype frequencies were calculated as the proportion of templates assigned to each clonotype. Shannon’s entropy was calculated as:

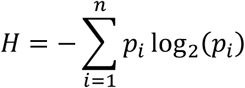

where (p_i_) represents the proportional abundance of clonotype (i) within the resampled repertoire. Richness was calculated within each bootstrap iteration as the total number of unique clonotypes detected in the resampled repertoire. S = number of clonotypes where ni > 0; where S represents richness and ni is the number of resampled templates assigned to clonotype i. Clonality was calculated as one minus Pielou’s evenness: *Clonality* = 1 – *J* where J is Pielou’s evenness index, calculated using Shannon’s entropy measures:

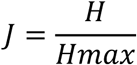

H_max_ represents the maximum possible diversity, assuming perfect evenness, and was calculated using richness measures: *Hmax* = ln(*S*)

### Deep-sequencing TCR longitudinal expansion analysis

Significantly expanded TCR clonotypes following CYT-101 and pembrolizumab were identified by comparing the screening time point (Screen) vs Visit 5 (VN5; prior to pembrolizumab infusion 1) and Visit 5 vs Visit 7-8 (post-pembrolizumab commencement) deep-sequencing data for each patient (n = 9). Productive TCR templates were aggregated by clonotype, and clonotypes with ≥5 templates in either time point were tested. Expansion was assessed using an exact binomial test, with the expected post-infusion probability adjusted for total productive template depth at each timepoint. Significance was corrected within each patient using the Benjamini–Hochberg multiple comparisons testing (P_adjusted_ < 0.01).

### Whole-blood immune monitoring

Phenotyping of blood cells was performed as previously described (27). Whole blood (50 µL) was transferred to a BD Trucount Tube (BD Biosciences cat# 663028), incubated with anti-CCR7–PE-Cy7 for 10 minutes, followed by LIVE/DEAD Fixable Near-IR and anti-CD45RA–FITC, anti-CD38– PerCP-Cy5.5, anti-CD45–APC, anti-CD4–R718, anti-CD56–BV421, anti-CD28–BV480, anti-CD3–BV711, anti-CD8 Super Bright 780, anti-CD16–PE, anti-CD27–PE-Dazzle 594, and anti-CD19–PE-Cy5. Samples were then incubated with BD FACS Lysing Solution (BD Biosciences RRID:AB_2868862) for 15 minutes and acquired using a BD LSRFortessa with FACSDiva software. Post-acquisition analysis was also performed using FlowJo software (BD Biosciences RRID:SCR_001456). A list of antibodies containing RRID numbers can be found in Supplementary Table 2.

### Functional immune monitoring using intracellular cytokine analysis

A multiparametric intracellular cytokine assay was used to assess changes in CMV-specific and global T-cell immunity following immunotherapy. T-cell responses were assessed at three time points: directly prior to the first CYT-101 infusion, prior to the final CYT-101 infusion, and throughout the course of pembrolizumab infusions. PBMC were stimulated with the custom CMV peptide pool used to generate CYT-101, or with Cell Stimulation Cocktail, a ready-to-use reagent mixture that contains the T cell mitogens, phorbol 12-myristate 13-acetate and ionomycin (Thermo Fisher Scientific eBioscience cat# 00-4970-93). Cells were incubated for 4 hours in the presence of anti-CD107a–FITC, GolgiPlug (BD Biosciences), and GolgiStop (BD Biosciences). Cells were washed and incubated with anti-CD4–Pacific Blue, anti-CD8–PerCP-Cy5.5 and LIVE/DEAD Near-IR. Cells were then washed, fixed and permeabilised with Cytofix/Cytoperm buffer, washed with Perm/Wash buffer, and incubated with anti-IFN-γ–Alexa Fluor 700, anti-IL-2–PE and anti-TNF–APC. Cells were washed with Perm/Wash buffer then acquired on a BD LSRFortessa with FACSDiva software. Post-acquisition analysis was performed using FlowJo software. A list of antibodies containing RRID numbers can be found in Supplementary Table 2.

### Immune checkpoint and regulatory T-cell analysis of patient PBMCs and CMV-specific T-cell products

Cryopreserved participant PBMC and matched CYT-101 CMV-specific T-cell products were thawed and counted before staining. For checkpoint phenotyping, cells were washed in FACS buffer (PBS supplemented with 2% FCS) and incubated with Fc receptor block for 10 minutes at room temperature. APC-conjugated peptide–HLA class I multimer selected according to the participant’s HLA type and product specificity was added for 10 minutes at room temperature. Anti-CCR7 antibody was added for a further 10 minutes, followed by incubation with antibodies against CD3, CD4, CD8, CD14, CD19, CD27, CD28, CD45RA, PD-1, LAG-3, TIGIT, and TIM-3, together with LIVE/DEAD Fixable near-IR Dead Cell Stain for 30 minutes. Cells were then washed and fixed in Cytofix fixation buffer. The multimers used recognised the HLA-A*01:01-restricted VTE and YSE peptides, HLA-A*02:01-restricted NLV, HLA-B*07:02-restricted TPR, HLA-B*08:01-restricted ELR, HLA-C*07:02-restricted CRV, or HLA-A*24:02-restricted QYD, as applicable. For regulatory T-cell analysis, patient PBMC were stained with a fixable LIVE/DEAD fixable near-IR Dead Cell Stain and antibodies against CD3, CD4, CD8, CD14, CD19, and CD25. Cells were fixed and permeabilised using TF Fix/PermBuffer (BD Biosciences) before staining for FOXP3. Cells were then washed in TF Perm/Wash Buffer (BD Biosciences) and fixed in Cytofix fixation buffer. Data were acquired on a BD LSRFortessa flow cytometerusing BD FACSDiva software, with post-acquisition analysis performed using FlowJo software. Viable CD14⁻CD19⁻CD3⁺ cells were gated for CD4⁺ and CD8⁺ T-cell analyses. Regulatory T cells were identified as CD4⁺CD25^high^FOXP3⁺ cells within the viable CD3⁺ T-cell population. A list of antibodies containing RRID numbers can be found in Supplementary Table 2.

## Data availability

Single cell RNAseq and TCR sequencing data generated in this study are available on the following link: 10.5281/zenodo.21800156. Raw FASTQ files will be deposited in the European Genome-Phenome Archive (EGA) pending institutional approval for data processing in line with the approved project. All other data are available in the article and its Supplementary files or from the corresponding author upon request. Source data are provided with this paper.

## Results

### Manufacturing and characterisation of allogeneic CMV-specific T cells

Four healthy CMV-seropositive donors were selected based on HLA type to generate allogeneic CMV-specific T-cell products (referred to as CYT-101). Peripheral blood mononuclear cells (PBMC) collected from a single blood donation from each donor were stimulated with a custom-designed pool of CMV-derived peptide epitopes (Supplementary Table 1), expanded for 14 days in the presence of IL-2, and cryopreserved at 4 × 10^7^ cells per vial. All four CYT-101 batches demonstrated high purity, comprising >79% CD3^+^ T cells (Supplementary Table 3). CD8^+^ T cells predominated in batches CYT-CMV-A001 and CYT-CMV-A002, whereas CYT-CMV-A003 and CYT-CMV-A006 contained a more balanced distribution of CD4^+^ and CD8^+^ T cells. Functional assessment demonstrated polyfunctional CMV-specific CD4^+^ and CD8^+^ T-cell responses in all four batches, characterised by cytokine production following antigen stimulation (Supplementary Figure 1A). CMV-specific T cells recognised epitopes presented by multiple HLA alleles (Supplementary Table 3) and exhibited potent cytolytic activity against HLA-matched target cells (Supplementary Figure 1B). Alloreactive potential was detected in CYT-CMV-A002 and CYT-CMV-A006 (Supplementary Table 3). These batches were excluded from use in participants with HLA alleles matching these potential alloreactivities.

To further define product composition, single-cell RNA sequencing (scRNA-seq) and paired T-cell receptor sequencing (scTCR-seq) were performed. Transcriptomic analysis revealed highly comparable gene-expression profiles across all four CYT-101 batches (Figure 1A)). Analysis of CMV-specific T cells identified using barcoded peptide–MHC multimers demonstrated a similar cellular architecture across batches, with CMV-specific populations enriched for proliferating, cycling, and terminal effector memory T-cell subsets (Figure 1B; Supplementary Figure 2). TCR repertoire analysis demonstrated broad clonotypic diversity (Supplementary Table 4), consistent with recognition of multiple CMV-derived epitopes and HLA restrictions, supporting the polyclonal nature of the CYT-101 product. All batches were dominated by highly enriched clonotypes (Figure 1C), and clonotypes were shared between T-cell clusters within each batch (Figure 1D). Characterisation of checkpoint marker expression in CYT-101 batches demonstrated consistent high levels of TIGIT, TIM-3 and LAG-3, particularly in CMV-specific T cells, and low PD-1 expression (Supplementary Figure 3A). Characterisation of costimulatory marker expression in CYT-101 batches demonstrated a high proportion of CMV-specific T cells expressing the CD28, and lower expression of CD27 (Supplementary Figure 3B).

**Figure 1:**
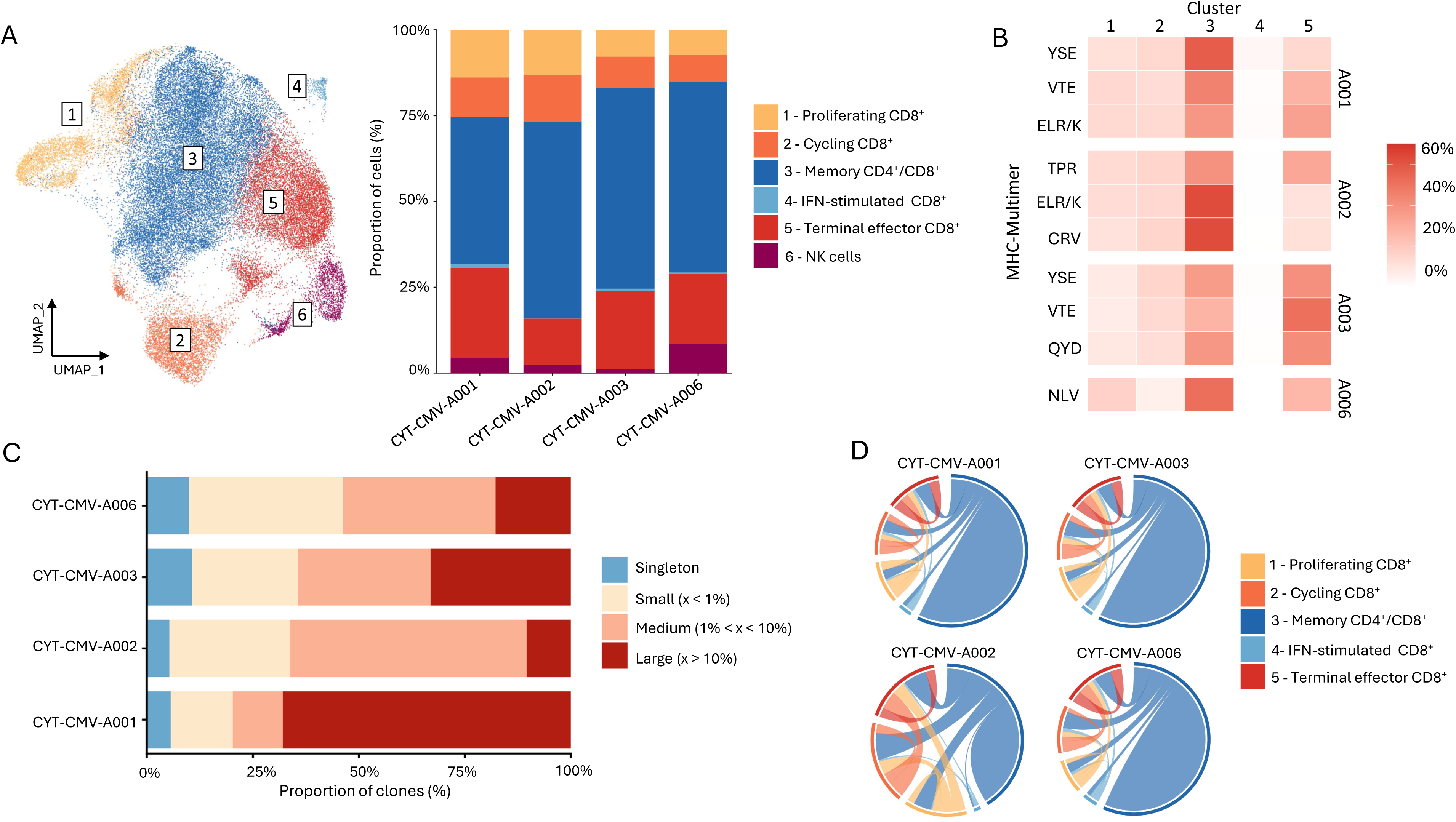
Single-cell characterisation of CYT-101 T cells. CMV-specific T cells from CYT-101 batches were sorted and analysed by scRNA sequencing. **(A)** UMAP projection of single-cell transcriptomes from four batches of CYT-101, coloured by cluster identity. Stacked bar graphs show the proportional cluster composition (%) of each batch. **(B)** Heatmap showing the distribution of multimer-specific T cells from each batch across UMAP-defined clusters. **(C)** Stacked bar graphs showing clonotype composition (%) within each batch, grouped by clone size category: singleton, small, medium, and large. Bars represent the proportion of clonotypes within each batch. **(D)** Chord diagrams showing clonotype sharing between clusters for each batch. Each segment represents a UMAP-defined cluster, and connecting ribbons indicate shared clonotypes between clusters. Ribbon thickness corresponds to the log_2_-transformed number of shared clones.

### Participant characteristics and treatment

This phase I study employed a standard 3+3 dose-escalation design to evaluate the safety and determine the maximum tolerated dose of CYT-101 in combination with pembrolizumab. Fourteen patients with recurrent GBM were screened for eligibility, of whom nine were enrolled and received study treatment (Figure 2). Four patients did not meet eligibility criteria: three had an Eastern Cooperative Oncology Group performance status of >1 and two did not have a HLA-matched batch of CYT-101. An additional patient was enrolled but subsequently withdrawn prior to treatment initiation due to clinical deterioration. Baseline clinical characteristics are summarised in Supplementary Table 5 and representativeness of study participants in Supplementary Table 6. Participants had a median age of 51 years (range, 39–62 years), 67% were male, and all had previously undergone surgical resection followed by radiotherapy and temozolomide-based chemotherapy (Supplementary Table 7). Patients received four weekly infusions of CYT-101 at dose levels of 2 × 10^7^, 4 × 10^7^, or 8 × 10^7^ cells per infusion. For each participant, a CYT-101 batch was selected based on matching of CMV-specific HLA restrictions (Supplementary Table 8). Six participants received CYT-CMV-A001, two received CYT-CMV-A002, and one received CYT-CMV-A003. Pembrolizumab treatment commenced one week after the final CYT-101 infusion, and all participants received at least one dose of pembrolizumab. Two participants (NEW-11 and NEW-12) had preliminary progressive disease detected at the completion of CYT-101 infusions and continued to receive one dose of pembrolizumab before discontinuation from trial treatment due to confirmed disease progression.

**Figure 2:**
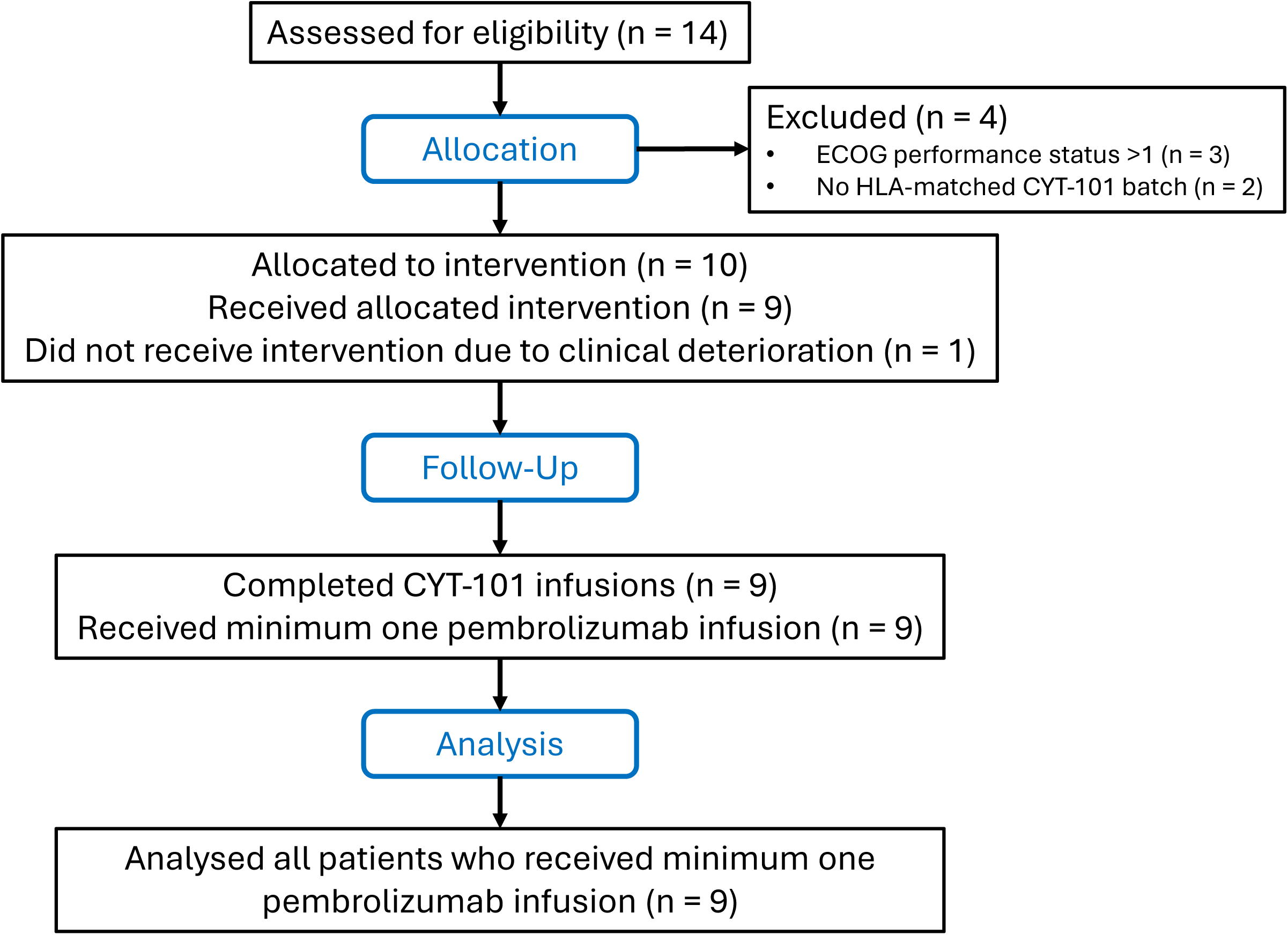
CONSORT diagram showing participant allocation, follow-up and analysis.

### Safety profile of CYT-101 combined with pembrolizumab

The primary endpoint was the incidence of dose-limiting toxicities (DLTs) within the 49-day DLT assessment period, extending to 21 days following the first pembrolizumab infusion. No DLTs were observed at any dose level, and the maximum tolerated dose was not reached. We therefore recommend the maximum administered dose for future exploration of allogeneic CMV-specific T cells as monotherapy or in combination with pembrolizumab. The combination of CYT-101 and pembrolizumab was generally well tolerated. Across the study, only six adverse events (AEs) were considered by the investigator to be possibly or definitely related to treatment (Supplementary Table 9): hypothyroidism (mild and moderate) and mild fever, maculo-papular rash and worsening fatigue. In total, 94 AEs were reported among the nine treated participants (Supplementary Table 10). Eleven serious adverse events (SAEs) occurred; however, all were assessed as unrelated or unlikely to be related to the study intervention. No treatment-related SAEs, cytokine release syndrome, immune effector cell-associated neurotoxicity syndrome, graft-versus-host disease, or unexpected safety signals were observed. Serial assessments of haematological, biochemical, coagulation, thyroid, and vital sign parameters revealed no clinically significant treatment-related abnormalities.

### Clinical outcomes following immunotherapy

Although the study was not powered to evaluate efficacy, exploratory analyses of clinical outcomes were performed as secondary endpoints. Participants were monitored for disease progression using magnetic resonance imaging (MRI) throughout the clinical trial. Representative MRI scans from four participants are shown in Figure 3. Clinical outcome for each participant based on mRANO criteria is provided in Supplementary Table 11. Tumour measurements for each participant based on mRANO are provided in Supplementary Table 12. Overall survival at 6 months (OS-6), defined as the proportion of participants alive 6 months after commencement of trial treatment, was 55.6% (95% CI: 20.4 to 80.5). Progression-free survival at 6 months (PFS-6), defined as the proportion of participants without disease progression based on mRANO criteria 6 months after commencement of trial treatment, was 11.1% (95% CI: 0.6 to 38.8). The median PFS was 2.1 months (Figure 4A), while the median overall survival was 9.1 months (Figure 4B). No complete or partial clinical responses were seen in this study. Of the nine treated participants, one had stable disease, five had progressive disease, and three were unevaluable by mRANO criteria. One participant, NEW-09 had a second surgery following progression (Figure 3) while four participants received bevacizumab following progression (Supplementary Table 7). Although most participants experienced disease progression during the study period, one participant (NEW-03), who had unmeasurable disease at baseline according to mRANO, remained free of radiological progression and clinical deterioration throughout the study period (Figure 4C). Two other participants (NEW-01 and NEW-06), who had unmeasurable disease at baseline according to mRANO, progressed during the study. Three participants (NEW-03, NEW-06, and NEW-10) remained alive at the time of the most recent participant follow-up (Figure 4C; Supplementary Table 11). While no definitive conclusions regarding efficacy can be drawn from this small phase I dose-escalation study, these findings provide preliminary evidence that administration of allogeneic CMV-specific T cells in combination with pembrolizumab has an acceptable safety profile and may warrant further evaluation in larger studies designed to assess clinical benefit.

**Figure 3:**
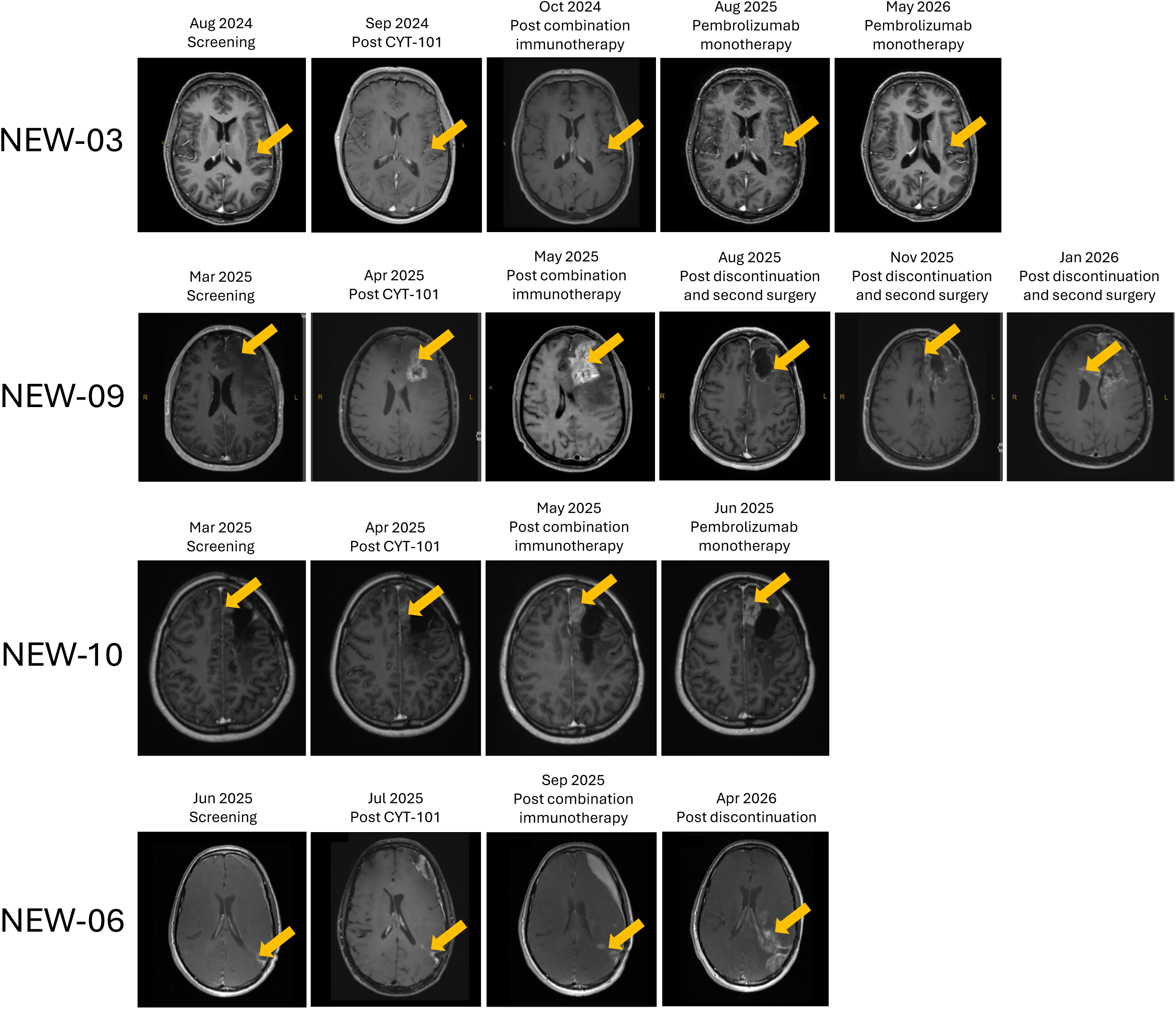
Representative MRI scans before and after combination immunotherapy. Sequential scans from participants NEW-06, NEW-09 and NEW-10, who experienced disease progression during pembrolizumab monotherapy and participant NEW-03, whose tumour did not progress during the clinical trial. Dates of each scan are shown. Details on time of progression for NEW-06, NEW-09 and NEW-10 can be found in Figure 4C.

**Figure 4:**
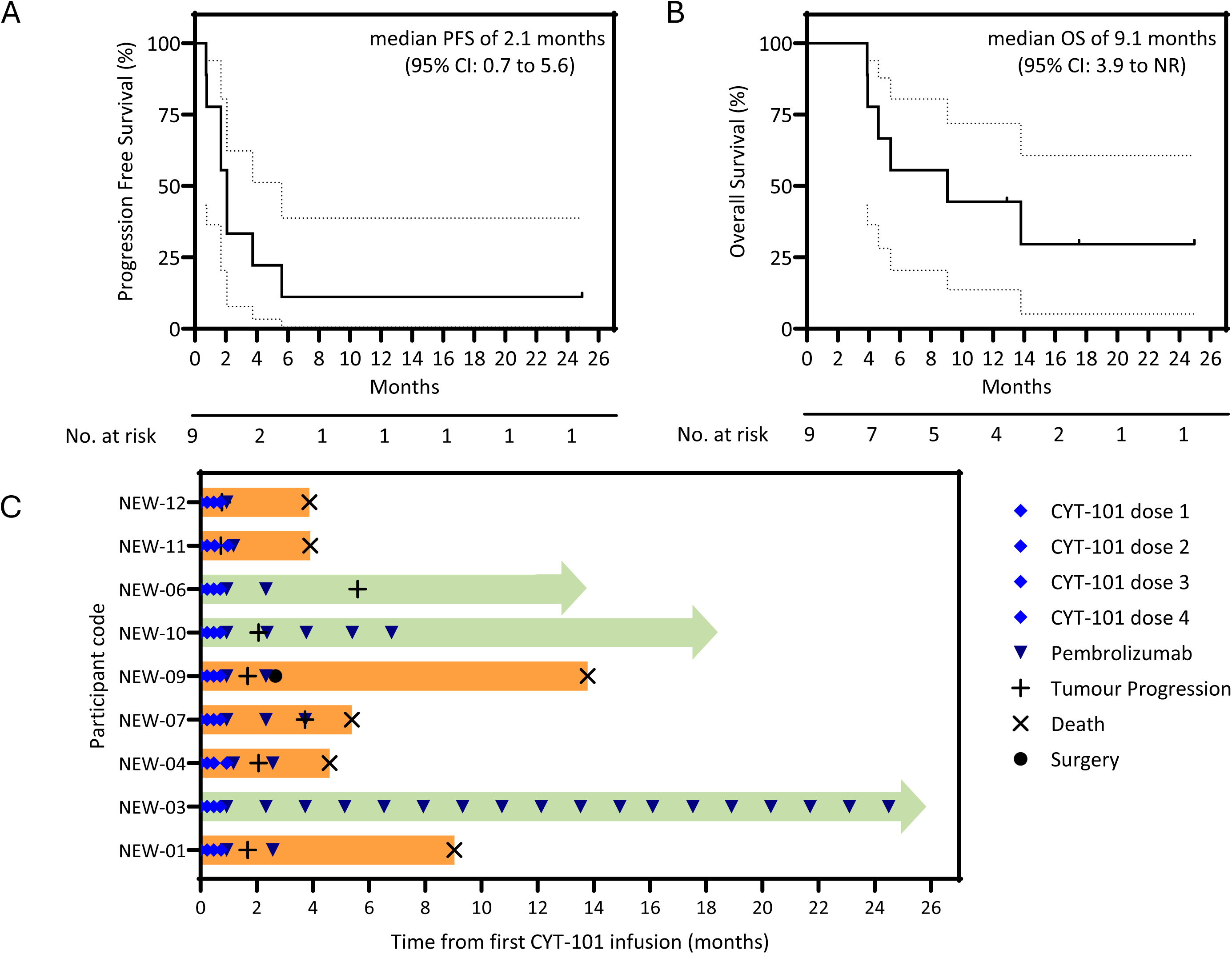
Clinical outcome following combination immunotherapy. **(A)** Progression-free survival from the start of treatment. Dotted lines represent 95% confidence intervals. **(B)** Overall survival from the start of treatment. Dotted lines represent 95% confidence intervals. **(C)** Swimmer plot of clinical course of individual participants, showing timing of CYT-101 and pembrolizumab infusions, tumour progression, death or surgery. Green bars represent participants who were still alive at last update; orange bars represent deceased participants. The key within the plot describes all symbols.

### Immune monitoring following immunotherapy

To evaluate the immunological effects of combination immunotherapy, peripheral blood samples were collected throughout the study. Analysis of immune cell subsets revealed no significant changes in the composition of the peripheral immune compartment following CYT-101 administration or subsequent pembrolizumab therapy (Supplementary Figure 4), indicating that treatment was not associated with broad perturbations of immune homeostasis. We next assessed CMV-specific immunity and global T-cell function at baseline, following completion of CYT-101 therapy, and after the initiation of pembrolizumab. No consistent changes were observed in the frequency or function of CMV-specific CD4^+^ or CD8^+^ T-cell responses across the cohort following either intervention (Figure 5, Supplementary Figure 5). In contrast, global T-cell function improved following CYT-101 administration, as evidenced by an increased proportion of CD4^+^ and CD8^+^ T cells producing IFN-γ in response to polyclonal stimulation (Figure 5). This was associated with significant differences in the proportion of polyfunctional CD4^+^ T cells (CD107a^-^ IFN-γ^+^IL-2^+^TNF^+^) and monofunctional CD4^+^ T cells (CD107a^-^IFN-γ^-^IL-2^+^TNF^-^) following CYT-101 therapy and after commencement of pembrolizumab (Supplementary Figure 5). These findings suggest that CYT-101 therapy was associated with enhanced peripheral T-cell functionality despite the absence of measurable expansion of circulating CMV-specific T-cell responses. Expression of the checkpoint markers PD-1, TIGIT, TIM-3 and LAG-3 did not change on CD8^+^ T cells, CD4^+^ T cells or CMV-specific T cells following CYT-101 therapy (Supplementary Figure 6). Expression of PD-1 appeared to decline after pembrolizumab treatment, although this is likely associated with pembrolizumab inhibition anti-PD-1 binding. The frequency of regulatory T cells didn’t demonstrate a consistent change following treatment (Supplementary Figure 7).

**Figure 5:**
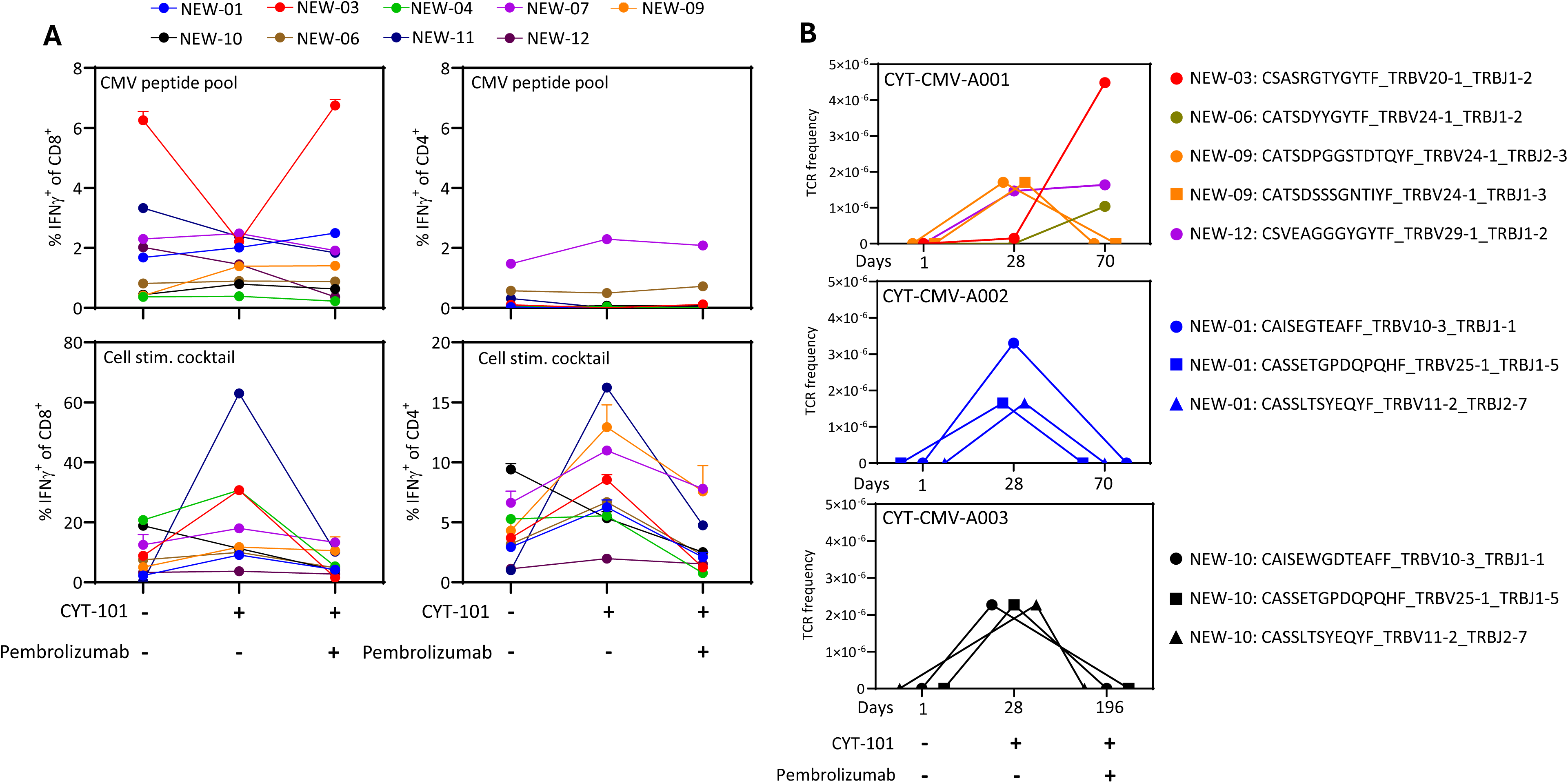
CMV-specific immune monitoring during combination immunotherapy. **(A)** PBMC isolated from participants before the first CYT-101 dose, after the final CYT-101 dose and prior to pembrolizumab, or during pembrolizumab treatment were assessed for IFN-γ-producing cells following stimulation with a pool of CMV peptides or a cell stimulation cocktail. Data represent the frequency of IFN-γ-producing CD8^+^ T cells (left panels) or IFN-γ-producing CD4^+^ T cells (right panels). (**B**) CYT-101 product-specific TCR clone frequencies in patient deep-sequencing data. Frequency of CYT-101 product-specific TCR clones detected by deep sequencing in participant samples (n = 9). Each dot represents a single CYT-101 product-specific TCR clone from an individual patient. Clones are grouped according to the CYT-101 batch received by each patient.

To investigate treatment-associated changes in the T-cell repertoire, genomic DNA isolated from PBMC collected at baseline, after CYT-101 therapy, and following pembrolizumab initiation was subjected to deep TCR β-chain (TRBV) sequencing using the ImmunoSEQ platform (Supplementary Table 13). Global repertoire analyses demonstrated no major alterations in overall TRBV architecture following either CYT-101 or pembrolizumab treatment (Supplementary Figure 8). However, interrogation of the dataset using TCR sequences identified in the CYT-101 products by single-cell TCR sequencing revealed the emergence of donor-derived CYT-101-associated clonotypes in six of nine participants after treatment initiation (Figure 6). In most participants, donor-associated clonotypes were detected following CYT-101 administration and subsequently declined after the commencement of pembrolizumab. In contrast, participants NEW-03, NEW-06, and NEW-12 demonstrated peak frequencies of donor-derived clonotypes after pembrolizumab initiation, suggesting that PD-1 blockade may have contributed to the persistence or expansion of transferred T-cell populations in a subset of patients.

**Figure 6.**
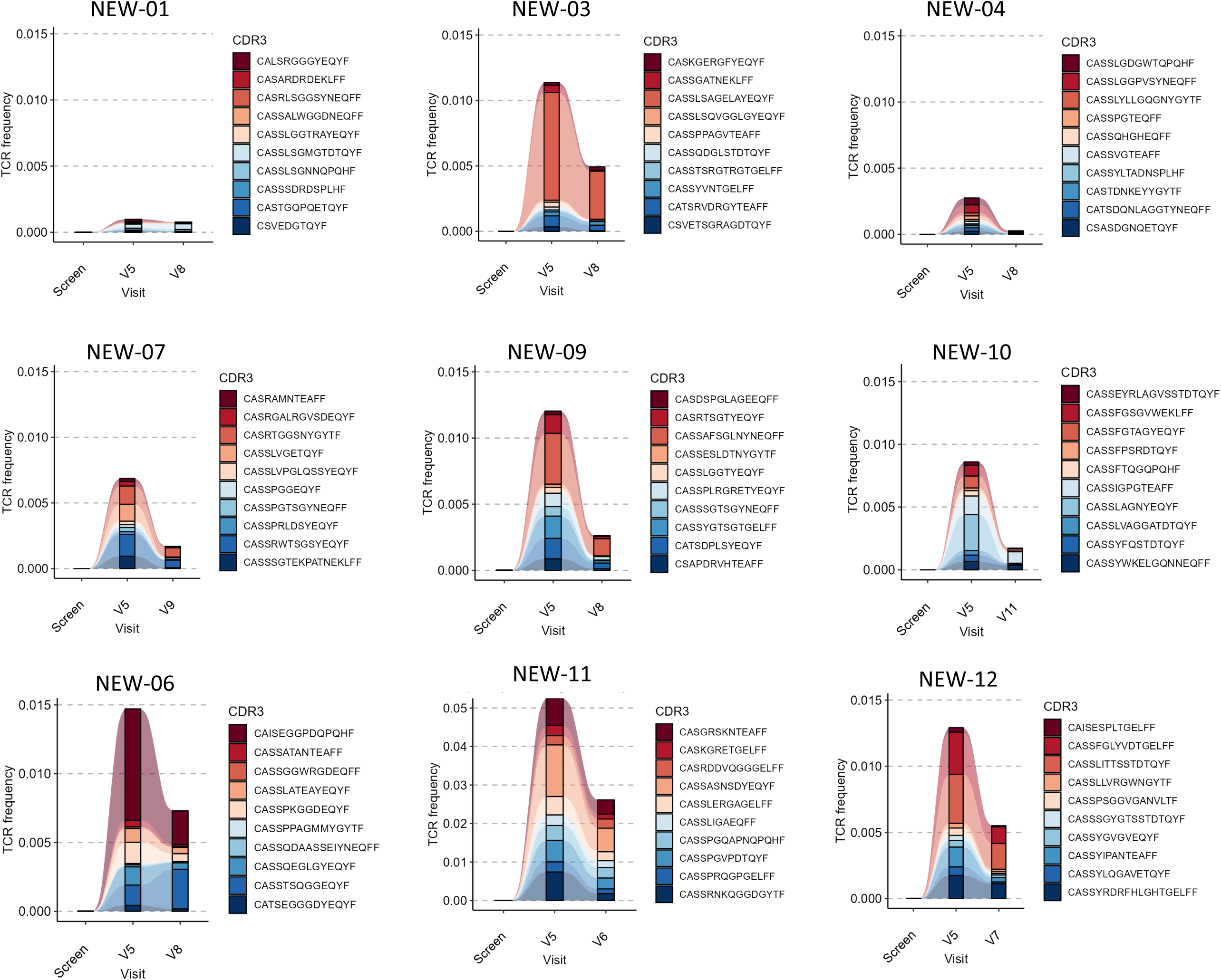
Longitudinal tracking of the top 10 expanded clonotypes post CYT-101 infusions. For each participant (n = 9), significantly expanded TCR clonotypes following all CYT-101 infusions were identified by comparing Visit 5 (prior to the first pembrolizumab infusion) with last visit (post-pembrolizumab) using an exact binomial test with Benjamini–Hochberg correction. Significantly expanded clonotypes were defined as enriched clonotypes post-pembrolizumab with P_adjusted_ < 0.01 and ≥5 productive templates detected post-infusion. The top 10 expanded clonotypes per participant, ranked by log2 fold change, were tracked across all available visits and visualised as alluvial plots using Immunarch (v0.10.3). Frequencies represent the proportion of productive TCR templates assigned to each clonotype at each visit.

To further characterise repertoire dynamics, differential abundance analysis was performed using a two-sided binomial test to identify significantly expanded TRBV sequences between sequential timepoints. All participants exhibited significant T-cell clonal expansion following CYT-101 therapy (Supplementary Table 14). The largest numbers of expanded clonotypes were observed in participants NEW-03 (n = 409) and NEW-11 (n = 381). Analysis of the ten most expanded clonotypes in each participant demonstrated a consistent pattern characterised by peak expansion following CYT-101 administration and subsequent contraction after pembrolizumab initiation (Figure 6), consistent with an acute response to adoptive T-cell transfer. In addition, a distinct population of significantly expanded clonotypes emerged following pembrolizumab treatment in five of nine participants (Supplementary Figure 9, Supplementary Table 15). Although the magnitude of expansion was generally lower than that observed following CYT-101 therapy, substantial repertoire remodelling was evident, particularly in participants NEW-09 (n = 152 expanded clonotypes) and NEW-03 (n = 148 expanded clonotypes). Three of the four participants who failed to demonstrate significant repertoire changes following pembrolizumab (NEW-07, NEW-11, and NEW-12) experienced relatively poor overall survival. Collectively, these findings demonstrate that CYT-101 adoptive immunotherapy induces measurable peripheral T-cell expansion and repertoire remodelling, while subsequent PD-1 blockade is associated with additional clonal dynamics in a subset of patients. The relationship between these immunological changes and clinical outcome warrants further investigation in larger studies.

## Discussion

GBM remains one of the most challenging malignancies to treat, with limited therapeutic advances despite substantial efforts to harness the immune system (28,29). In this phase I study, we evaluated the safety and biological activity of CYT-101, an allogeneic CMV-specific T-cell therapy, administered in combination with pembrolizumab in patients with recurrent GBM. The combination was well tolerated, with no dose-limiting toxicities, treatment-related serious adverse events, graft-versus-host disease, cytokine release syndrome, or neurotoxicity observed at any dose level. Importantly, longitudinal immune monitoring demonstrated evidence of allogeneic donor-derived T-cell persistence and treatment-associated remodelling of the peripheral T-cell repertoire, supporting the immunological activity of this combination approach.

Immune checkpoint blockade has transformed the treatment of several solid tumours; however, clinical trials of anti-PD-1 therapy in GBM have failed to demonstrate meaningful improvements in survival (30–32). One explanation for this lack of efficacy is the profoundly immunosuppressive and immunologically “cold” nature of the GBM microenvironment, which is characterised by limited T-cell infiltration, extensive myeloid-mediated immune suppression, T-cell dysfunction, and multiple mechanisms of immune evasion (33,34). Checkpoint blockade alone may therefore be insufficient in the absence of a pre-existing tumour-reactive T-cell population (35,36). We previously demonstrated that tumour-infiltrating CMV-specific T cells express elevated levels of PD-1 following adoptive transfer, suggesting that transferred T cells may become susceptible to checkpoint-mediated inhibition within the tumour microenvironment (23). The current study was designed to address this limitation by combining adoptive cellular therapy with PD-1 blockade. The favourable safety profile observed supports further clinical evaluation of this strategy.

A notable feature of this study was the use of an allogeneic virus-specific T-cell product. Unlike autologous approaches, allogeneic CYT-101 could be manufactured in advance, cryopreserved, and administered without delay. This approach overcomes many logistical barriers associated with individualised cell therapies, including manufacturing failures, variable product quality, and prolonged vein-to-vein times. Importantly, no evidence of graft-versus-host disease was observed, despite repeated infusions of partially HLA-matched donor T cells. These findings are consistent with our previous studies demonstrating the safety of virus-specific T-cell therapies in patients with cancer and transplant recipients (22,37–40) and support the feasibility of developing off-the-shelf cellular immunotherapies for patients with GBM.

Although this study was not designed to evaluate efficacy, several observations suggest that CYT-101 induced measurable immunological activity. Following therapy, we observed enhanced global CD4^+^ and CD8^+^ T-cell function, as demonstrated by increased IFN-γ production after polyclonal stimulation. Interestingly, these changes occurred in the absence of consistent expansion of circulating CMV-specific T cells. This finding suggests that the immunological effects of CYT-101 may extend beyond simple expansion of virus-specific populations and may involve broader activation of endogenous immune responses. Similar phenomena have been observed in other cellular therapy settings, where adoptively transferred T cells promote epitope spreading and secondary immune activation (22,41,42).

One of the most important findings of this study was the detection of donor-derived CYT-101 TCR clonotypes in six of nine participants following treatment. These data provide direct evidence that transferred allogeneic T cells can persist in vivo following infusion. The ability to track infused clonotypes using single-cell TCR sequencing and bulk repertoire analysis has emerged as an important tool for understanding the fate of adoptively transferred cells and their contribution to therapeutic responses (43,44). While donor-derived clonotypes generally contracted over time, a subset of patients demonstrated peak frequencies following the commencement of pembrolizumab, raising the possibility that PD-1 blockade may enhance the persistence or expansion of transferred T cells. Previous preclinical and clinical studies have shown that checkpoint blockade can improve the survival, expansion, and functional capacity of adoptively transferred T cells (45,46). Although this observation requires validation in larger studies, it provides preliminary support for the biological rationale underpinning this combination strategy.

Deep TCR repertoire sequencing further revealed substantial remodelling of the peripheral T-cell compartment following treatment. All participants exhibited significant clonal expansion after CYT-101 administration, while a distinct wave of repertoire changes emerged following pembrolizumab treatment in a subset of patients. These findings suggest that the two therapies may exert complementary immunological effects. CYT-101 appeared to drive an initial burst of T-cell expansion, whereas pembrolizumab was associated with subsequent diversification and remodelling of the T-cell repertoire. Similar treatment-associated repertoire changes have been associated with clinical responses to adoptive cellular therapies and immune checkpoint blockade in other cancers (47,48). Notably, the participants demonstrating the most pronounced repertoire dynamics included those with prolonged survival, suggesting that treatment-induced immune remodelling may represent a biomarker of therapeutic activity. However, the small sample size precludes definitive conclusions regarding associations between repertoire changes and clinical outcomes.

The findings of this phase I study should be interpreted in light of several limitations. As a phase I dose-escalation trial, the primary objective was safety rather than efficacy, and the small cohort size limited statistical power for clinical and correlative analyses. Furthermore, the contribution of CYT-101 cannot be separated from the effect of pembrolizumab, and immune monitoring was performed primarily in peripheral blood, which may not fully capture immune dynamics occurring within the tumour microenvironment. Future clinical studies using this treatment strategy should include a cohort of participants receiving CYT-101 alone, in addition to a cohort receiving the combined therapy. Future studies incorporating paired tumour biopsies, cerebrospinal fluid analyses, and spatial immune profiling will be important to determine whether donor-derived T cells traffic to tumour sites and mediate local immune remodelling. In addition, although CMV-associated antigens have been reported in GBM (49–51), the extent and biological significance of CMV expression in gliomas remains an area of ongoing investigation (13,19,52,53).

In conclusion, this study demonstrates that allogeneic CMV-specific T-cell therapy can be safely combined with PD-1 blockade in patients with recurrent GBM. The detection of donor-derived clonotypes, enhancement of T-cell function, and extensive treatment-associated remodelling of the T-cell repertoire provide evidence of immunological activity and support further evaluation of this approach. More broadly, these findings suggest that adoptive virus-specific T-cell therapy may provide a strategy to increase tumour-reactive immunity and potentially overcome barriers that have limited the efficacy of immune checkpoint blockade in GBM.

## Data Availability

All data produced in the present study are available upon reasonable request to the authors.

https://10.5281/zenodo.21800156

## Author contributions

RK and DW conceived and designed the clinical trial. PM, PK, AP, LB, SF, JR, MS, MV and CS contributed to CYT-101 manufacture, immunomonitoring and data analysis. MAN and HR contributed to essential document drafting, trial approvals, and data and record management. JK and VB contributed to patient recruitment and management, data collection and record management. DW, HA and AS, contributed to patient recruitment, treatment and medical oversight. CS, RK, PM and DW wrote the original draft of the manuscript and all authors contributed to the manuscript review and final drafting.

## Funding support

This clinical trial was supported by a Medical Research Future Fund CUREator Grant to Cyteph Pty Ltd. This clinical trial was also supported in part by a research grant from the Investigator-Initiated Studies Program of Merck Sharp & Dohme (Australia) Pty Ltd. The opinions expressed in this paper are those of the authors and do not necessarily represent those of Merck Sharp & Dohme (Australia) Pty Ltd. This work was also supported in part by a generous donation from the William and Hilde Chenhall Research Trust.

## Acknowledgments

We acknowledge Stem Cell Donors Australia for assistance with selection and recruitment of blood donors for CYT-101 manufacture. We would like to acknowledge logistical support and quality management provided by Q-Gen Cell Therapeutics staff for manufacturing of CYT-101 T-cell therapies. We would like to that Dr Gunter Hartel for support with the study statistical design and Sally-Ann Pearson for help with trial coordination. We would like to thank Dr Dalveer Singh for his independent MRI reviews and mRANO reporting. We also acknowledge the staff at the Wesley Research Institute, Wesley Hospital and Sullivan Nicolaides Pathology for their support of the trial

## Conflict of Interest Disclosures

PM, CS and RK have scientific leadership roles at Cyteph Pty Ltd. CS and RK are listed as inventors on patents and patent applications relevant to this study.

## Notes

### Clinical Trial

NCT06157541

### Author Declarations

Ethical approval for the cell therapy manufacturing project was obtained from the Lifeblood Human Research Ethics Committee (HREC) and the QIMR Berghofer Medical Research Institute HREC. The clinical trial was reviewed and approved by the Metro North Health HREC and the UnitingCare Health HREC.

